# Sodium Valproate Modulates Cortical Morphology in Juvenile Myoclonic Epilepsy

**DOI:** 10.1101/2024.01.09.24301048

**Authors:** Bernardo Crespo Pimentel, Giorgi Kuchukhidze, Fenglai Xiao, Lorenzo Caciagli, Julia Höfler, Lucas Rainer, Martin Kronbichler, Christian Vollmar, John S Duncan, Eugen Trinka, Matthias Koepp, Britta Wandschneider

## Abstract

**Importance:** Idiopathic generalized epilepsy syndromes in are associated with cortical thinning of the premotor areas. Whether this represents an underlying disease signature, a consequence of seizure activity or is related to anti-seizure medication is unknown.

**Objective:** To investigate valproate-related effects on cortical morphology in people with juvenile myoclonic epilepsy (JME), one of the most common IGE syndromes.

**Design:** Retrospective neuroimaging case-control study.

**Setting:** Recruitment took place in epilepsy clinics in two European epilepsy referral centers.

**Participants:** We matched individuals with JME on valproate (*n*=36) to a group of healthy controls (*n*=36), as well as individuals with JME not on valproate (*n*=36) and a group of people with temporal lobe epilepsy (*n*=19) on valproate using propensity scores for age, sex, occurrence of bilateral tonic-clonic seizures and drug refractoriness.

**Main outcomes and Measures:** All participants underwent structural T1-weighted brain imaging and MRI-derived vertex-wise measurements of cortical thickness were calculated. Results were reported in effect-sizes using cohen’s *d*.

**Results:** Compared to healthy controls, individuals with JME on valproate demonstrated cortical thinning bilaterally in the precentral gyri (left: *d* = -1.0, *p* <.001; right: d = -1.0, *p* <.001). This effect was localized to the left precentral gyrus when comparing JME on VPA with individuals with JME not on valproate (*d* = -0.89; *p*<.01) or with individuals with temporal lobe epilepsy on valproate (*d = -0*.*18, p* <.001). No significant differences in cortical thickness were detected between individuals with JME not on valproate and healthy controls. Cortical thinning in precentral gyrus, postcentral gyrus and superior frontal was more marked with increasing valproate dose (left: *t* = -6,65, *p* <.0001; right: *t* = -5,20, *p* <.0001).

**Conclusions and Relevance:** In this cross-sectional study of 91 patients with epilepsy, valproate treatment was associated with JME-specific and dose-dependent cortical thinning of the precentral gyri. Our findings suggest a valproate-induced reorganization of disease-specific areas in JME that may not only help explain the high efficacy of this medication in IGEs but also potentially account for previously described changes of cortical thickness in this disease group.

## Introduction

Sodium valproate (VPA) is a widely used anti-seizure medication (ASM), which is highly effective in idiopathic generalized epilepsies (IGEs).^1^ The mechanisms behind this remain largely unknown. Novel insights keep unraveling valproate’s teratogenic effects and its association with abnormal neurocognitive development if exposed in-utero, thereby limiting its usage.^2,3^ Juvenile myoclonic epilepsy (JME) is a common form of IGEs and neuroimaging studies have revealed gray matter loss in the precentral and medial prefrontal areas as a syndrome-specific feature.^4,5^ Whether these changes reflect an underlying disease signature, consequence of ongoing seizure activity or drug-associated effects, is unknown. Here, we aimed to investigate VPA-related effects on cortical morphology in JME and test for syndrome-specificity. First, we analyzed a neuroimaging dataset of people diagnosed with JME on VPA and compared it to healthy controls and to a JME group not on VPA. Then, to assess syndrome specificity, we compared the JME group on VPA with a temporal lobe epilepsy (TLE) group also on VPA.

## Methods

### Participants

We analyzed imaging data obtained between 2009 and 2017 of people with JME, TLE and healthy controls recruited from epilepsy clinics at the University College London Hospitals (UCLH), London, United Kingdom and the Christian Doppler University Hospital in Salzburg, Austria (CDK). All participants had high-resolution T1-weighted MRI scans. We excluded individuals with insufficient image quality and brain lesions other than hippocampal sclerosis in the TLE subgroup. Individual ASM load was calculated as previously reported.^6^ The study was approved by the ethics committee of UCLH and Salzburg state. All participants provided written informed consent. We followed the recommendations of the “Strengthening the Reporting of Observational Studies in Epidemiology” (STROBE) reporting guidelines.

### Image Preprocessing

The MRI acquisition protocols are described in eAppendix 1 in the Supplement. Cortical thickness was estimated vertex-wise through a surface-based framework using FreeSurfer and measurements were subsequently down-sampled to a common surface template (preprocessing pipeline described in eAppendix 2, Supplement). Prior to statistical analysis, data were adjusted for scanner-related batch effects using the validated Combat Tool for Harmonization of Multi-Site Imaging Data in R.^7^

### Statistical Analysis

To enable group comparison, we adopted a nearest-neighbor propensity score matching using age, sex, occurrence of generalized tonic-clonic seizures (GTCS) and drug resistance to calculate the propensity scores. Demographic and clinical data were analyzed with R studio (v2023.03.0+386). Kruskal-Wallis test was used to compare continuous clinical characteristics between subgroups. Pearson’s Chi-Square was used for categorical data. Vertexwise cortical thickness measurements were analyzed with BrainStat for Matlab.^8^ Fitted linear models for in-between JME group comparisons were corrected for a fixed effect of age, sex and group, whereas models for comparisons with TLE were additionally corrected for total ASM load. Models for comparison with healthy controls were corrected for age and sex. We report effect-sizes using Cohen’s *d* considered significant after correction for multiple comparisons using random field theory^9^ at family-wise error (FWE) <.05.

## Results

After propensity score matching, we generated four groups: i) JME on VPA (*n*=36), ii) JME not on VPA (*n*=36), iii) TLE on VPA (*n*=19) and iv) healthy controls (*n*=36). All disease groups were comparable for age, sex, disease duration, presence of GTCS and drug resistant disease. The TLE on VPA group had a higher total ASM load than both JME groups (TLE on VPA 2.37 [1.43, 3.27] vs. JME on VPA 1.20 [0.67, 1.77] vs JME not on VPA 1.00 [0.46, 1.37], respectively; Kruskal-Wallis T = 10.11, *p* <.001).

### Effect of VPA use

There was no significant differences in cortical thickness (see eFigure 1 in Supplement) between all individuals with JME and healthy controls (see eFigure 1 in Supplement). Compared to healthy controls, JME on VPA showed bilateral cortical thinning within the precentral gyri (left and right: *d* = -1.0, *p*_FWE_ <.001) and thickening of the right posterior cingulum and lingual gyrus (*d* = 0.71, *p*_FWE_ <.0001). Compared to individuals with JME not on VPA, the JME on VPA group showed cortical thinning in the left precentral gyrus (*d* = -0.89; *p*_FWE_ <.01) and thickening in the posterior cingulum bilaterally (left: *d* = 0.59, *p*_FWE_ <.0001; right: *d* = 0.61, *p*_FWE_ <.0001).

Compared to TLE on VPA, JME on VPA showed slight grey matter atrophy in the left precentral gyrus and parts of the left middle frontal gyrus (*d = -0*.*18, p*_FWE_ <.001) as well as bilaterally in both parahippocampal gyri (left: *d* = -0.27, *p*_FDR_ <.001; right: *d* = -0.46, *p*_FDR_ <.0001). No significant differences were seen between JME not on VPA and healthy controls. Results are displayed in Figure 1A-D and provided in detail in eTable 1 in Supplement.

**Figure 1.**
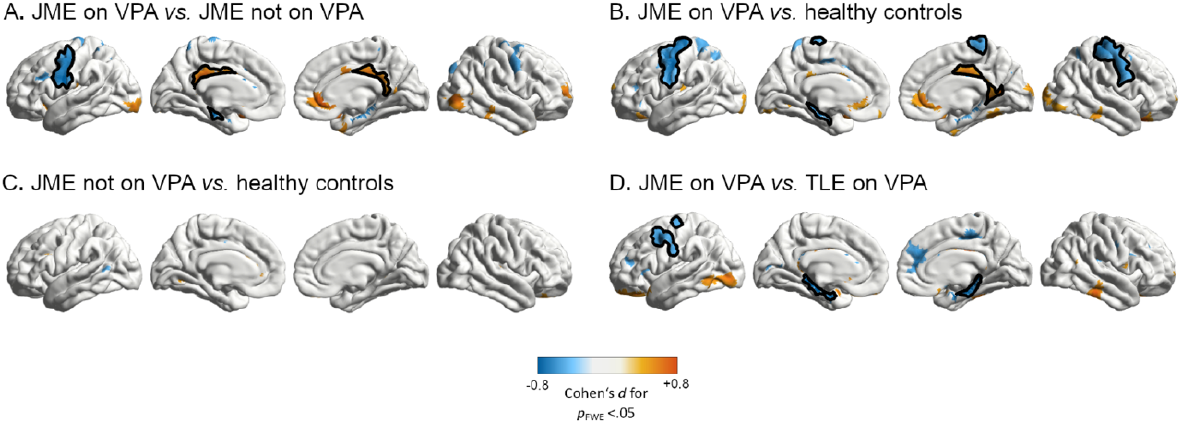
Effect of VPA across groups. Mass univariate analysis showing group comparisons of corticalthickness between individuals with JME on VPA, JME off VPA, TLE on VPA and healthy controls. The JME group on VPA shows significant cortical thickness deficits in the left precentral gyrus and a thickened posterior cingulum when compared to the JME group not taking VPA (A) as well as to healthy controls (B). Similar, although less extensive cortical thinning comprising the left precentral gyrus is seen on the JME on VPA group when compared to the TLE on VPA group (D), suggesting effect-specificity to JME. No significant structural abnormalities were detected when comparing the JME off VPA group with healthy controls (C), thereby controlling for disease-related effects. Clusters are color-coded according to the corresponding effect size estimates as reported by Cohen’s *d* (see color bar). Clusters that survived multiple comparisons correction using random field theory at *p*_FWE_ <.05 were manually outlined in black.

**Figure 2.**
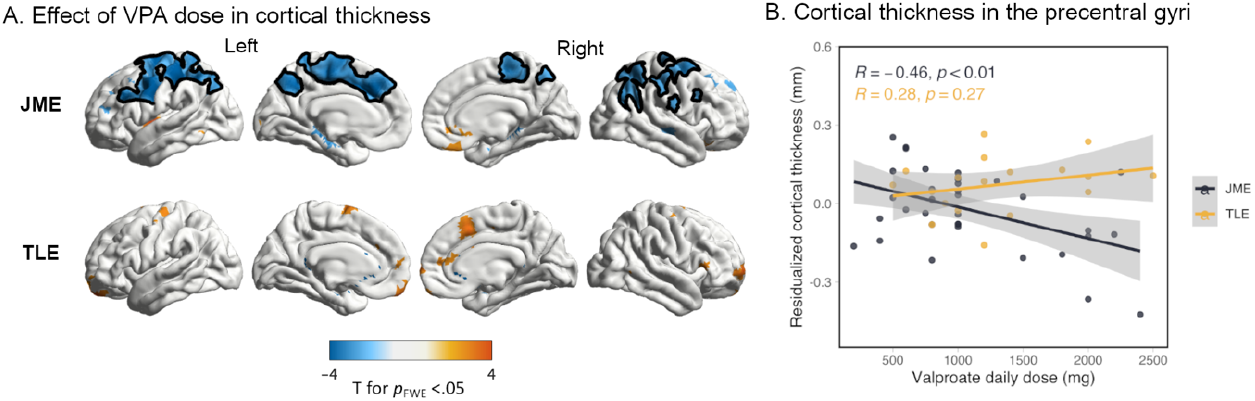
VPA dose and cortical thickness across JME and TLE. In (A), vertexwise linear regression T-map shows changes in cortical thickness as predicted by sodium valproate (VPA) dose in juvenile myoclonic epilepsy (JME) and temporal lobe epilepsy (TLE) after adjusting for total anti-seizure medication (ASM) load, age and sex. Clusters are color-coded according to the corresponding T statistic (see color bar). Clusters that survived multiple comparisons correction using random field theory at *p*_FWE_ <.05 were manually outlined in black. (B) shows linear regression scatterplots illustrating the association between VPA dose and cortical thickness specifically in the bilateral precentral gyri. Individualized cortical thickness values were extracted from parcellations of the Destrieux brain atlas incorporated in Freesurfer corresponding to the left and right precentral gyrus. Cortical thickness values were averaged across both parcels and adjusted for age, sex and total ASM load via multiple regression. Displayed are the residualized cortical thickness values plotted against the VPA dose for JME and TLE patients.

### Effect of VPA dose

In those with JME on VPA, cortical thinning correlated with increasing doses of VPA and affected the pre- and postcentral gyri, the superior frontal gyrus and precuneus in on both hemispheres (left: *t* = -6,65, *p*_FWE_ <.0001; right: *t* = -5,20, *p*_FWE_ <.0001). No significant effect was seen in the TLE group on VPA. Results are displayed in Figure 1B and provided in detail in eTable 2 in Supplement.

## Discussion

We found an association between VPA treatment and cortical thinning of the motor areas in individuals with JME. Gray matter atrophy has been described in the context of VPA treatment, but the regions affected have not been consistent across studies.^10-12^ In our study, whereas cortical thinning in JME on VPA affected the precentral gyri when compared to JME not on VPA, healthy controls, and, to a lesser extent, TLE on VPA, no changes were seen in those with JME not taking VPA. This suggests that VPA is a key modulator of cortical morphology in disease-specific regions and its effect is specific to JME. Our hypothesis is further corroborated by a dose-dependent effect of VPA on cortical thinning of the motor regions. In this way, our findings suggest that the results of a recent large-scale MRI study which demonstrated gray matter atrophy in IGE syndromes localized to the precentral gyri may be attributable to some extent to VPA.^4^

The motor areas are major hubs in the pathophysiology of IGE and functional MRI studies have identified hyperactivation patterns of the motor cortices which not only likely facilitate seizure generation but also normalize with increasing VPA dose.^13-15^ We thus hypothesize that VPA-associated structural reorganization of the motor cortices is related to previoulsy described network-stabilizing effects of VPA in the precentral gyri.^14^ However, to further corroborate this notion, our findings need to be correlated with disease activity.

Limitations include the relatively small group sizes, lack of information regarding time duration of VPA treatment, neuropsychological testing and adequate markers of disease activity. Furthermore, the JME group not on VPA also included individuals who had previously been on VPA, thereby restricting group homogeneity. As many participants have been treated with VPA in the past, we hypothesize that the observed grey matter effects of VPA might be reversible. Our findings raise questions about the mechanisms behind VPA efficacy in IGE syndromes and the pathophysiology of JME, while highlighting the need to control for ASM-related effects in cortical morphology studies.

**Table 1.** Demographic and clinical characteristics.

|  | Controls (n = 36) | JME on VPA<br>(n = 36) | JME not on VPA<br>(n = 36) | TLE on VPA<br>(n = 19) | Test<br>statistic | *P* value | Post-hoc *P* values |
| --- | --- | --- | --- | --- | --- | --- | --- |
| Age at time of scan,<br>years (IQR) | 32 [25, 37] | 28 [24, 32] | 32 [25, 39] | 34 [30, 44] | 1.67 | .103 |  |
| Female, No. (%) | 18 (50) | 22 (61) | 14 (39) | 11 (58) | 1.13 | .266 |  |
| Drug resistant, No. (%) | NA | 17 (47) | 19 (53) | 14 (74) | 1.48 | .163 |  |
| GTCS, No. (%) | NA | 9 (25) | 13 (36) | 9 (47) | 0.27 | .246 |  |
| Disease duration, years<br>(IQR) | NA | 14 [10, 20] | 16 [11, 28] | 16 [8, 29] | 0.25 | .434 |  |
| Total ASM load at<br>scanning, n (IQR) | NA | 1.00 [0.46, 1.37] | 1.20 [0.67, 1.77] | 2.37 [1.43,<br>3.27] | 10.11 | <.001 | JME VPA+/JME VPA-: .211<br>JME VPA+/TLE VPA-: <.05 |

Abbreviations: JME, juvenile myoclonic epilepsy; VPA+, individuals on sodium valproate; VPA-, individuals off valproate; TLE, temporal lobe epilepsy; IQR, interquartile range; GTCS, generalized tonic-clonic seizures;

## Data Availability

Data produced in the present study are available upon reasonable request to the authors.

