## Supplement for "Sodium Valproate Modulates Cortical Morphology in Juvenile Myoclonic Epilepsy"

Supplemental Online Content

**eAppendix 1.** MRI acquisition protocol in participating centers

**eAppendix 2**. MRI preprocessing pipeline

**eFigure 1**. Comparison of cortical markers between individuals with JME vs. healthy controls

**eTable 1:** Clusters that survived familywise-error correction in group comparisons for cortical thickness

**eTable 2:** Clusters that survived familywise-error correction in effect of VPA dose for cortical thickness

eAppendix 1: MRI Acquisition Protocol in Participating Centers

Structural MRI data at Paracelsus Medical University (PMU) were obtained with a Siemens Magnetom Prisma-fit 3T scanner with a 64-channel head/coil. High-resolution T1-weighed images were acquired using a 3D multiecho MPRAGE sequence (repetition time 2.4ms, echo time 2.2ms, inversion time 1060ms, flip angle = 8°, matrix = 320x300, voxel size = 0.8x0.8x0.8mm^3^). T1-weighted structural MRI data at University College London (UCL) were obtained on a 3T General Electric (Milwaukee, MN) using a 3D fast spoiled gradient echo sequence (repetition time 7.2ms, echo time 2.8ms, inversion time 450ms, flip angle = 20°, matrix = 256x256, voxel size = 1.1x1.1x1.1mm^3^).

eAppendix 2. MRI Preprocessing Pipeline

MRI datasets were processed using the standardized Freesurfer processing pipeline. ^1^ The processing stream consisted of automated transformation to Talairach space, skull stripping, intensity normalization and segmentation of white/grey matter tissue, resulting in extraction of surface meshes composed of approximately 150,00 vertices in each hemisphere. Surface extractions were visually assessed and manually corrected. Individual surfaces were then registered to an average template surface, composed of 20484 vertices and metric maps were smoothed with a smoothing kernel of 20 mm at FWMH.

eFigure 1: *Comparison of cortical markers between individuals with JME vs. healthy controls*

A. Individuals with JME (n=72) *vs.* healthy controls (n=36)

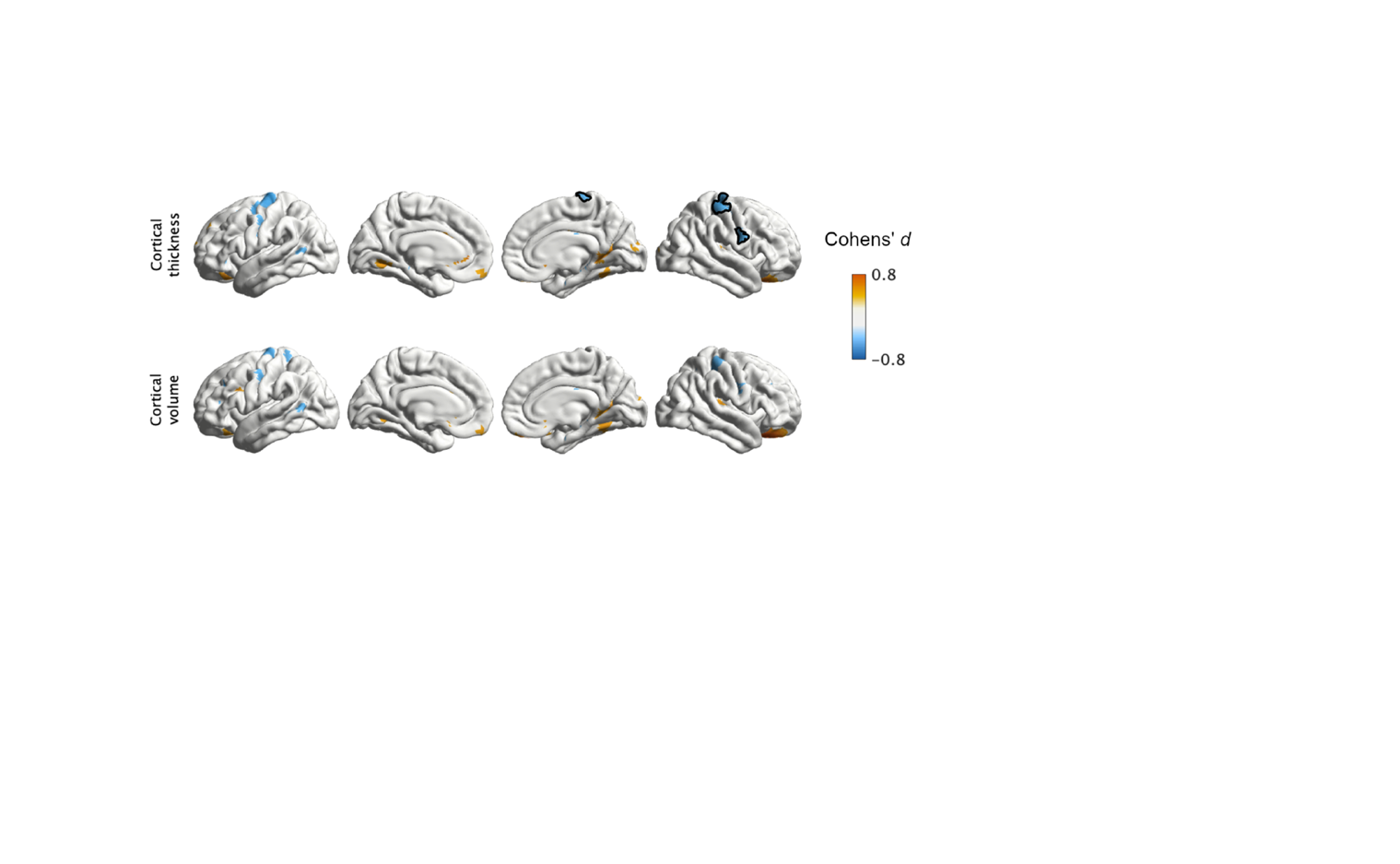

Univariate group analysis shows how cortical thickness and volume differ across individuals with JME vs healthy controls (syndrome-related effects). Both multiple regression models were covaried for age and sex. We report effect sizes with Cohens’*d* for areas with cluster-uncorrected *p*-values <0.05. Significant clusters corrected for multiple comparisons using random field theory (FWE) at *p*_FWE_ <0.05 are outlined in black.

| **eTable 1:** *Clusters that survived familywise-error correction in group comparisons for cortical thickness* | | | |
| --- | --- | --- | --- |
| ***JME vs. healthy controls*** | | | |
| **Cluster** | **Cohens *d* (95% CI)** | ***p***_FWE_ | **Regions** |
| 1 | -0.65 (-1.2, -0.41) | <.05 | Right superior precentral gyrus |
| ***JME on VPA vs. JME not on VPA*** | | | |
| **Cluster** | **Cohens *d* (95% CI)** | ***p***_FWE_ | **Regions** |
| 1 | -0.89 (-1.4, -0.40) | <.01 | Left precentral gyrus |
| 2 | 0.61 (0.13, 1.1) | <.0001 | Right posterior cingulum |
| 3 | 0.59 (0.11, 1.1) | <.0001 | Left posterior cingulum |
| 4 | 0.07 (-0.39, 0.54 | <.01 | Right medial parahippocampal gyrus |
| 5 | -0.09 (-0.56, 0.37) | <.01 | Left medial parahippocampal gyrus |
| ***JME on VPA vs. healthy controls*** | | | |
| **Cluster** | **Cohens *d* (95% CI)** | ***p***_FWE_ | **Regions** |
| 1 | 0.71 (0.23, 1.2) | <.0001 | Right posterior cingulum, right lingual gyrus |
| 2 | -1.0 (-1.5, -0.54) | <.001 | Left precentral gyrus, left posterior superior frontal gyrus |
| 3 | -1.0 (-1.5, -0.50) | <.001 | Right precentral gyrus, right postcentral gyrus |
| 4 | 0.01 (-0.48, 0.45) | <.05 | Left medial parahippocampal gyrus |
| ***JME on VPA vs. TLE on VPA*** | | | |
| **Cluster** | **Cohens *d* (95% CI)** | ***p***_FWE_ | **Regions** |
| 1 | -0.46 (-1.1, 0.12) | <.0001 | Right parahippocampal gyrus |
| 2 | -0.27 (-0.85, 0.31) | <.001 | Left parahippocampal gyrus |
| 3 | -0.18 (-0.77, 0.40) | <.001 | Left precentral gyrus, left middle frontal gyrus, left inferior postcentral gyrus |
| Abbreviations: JME, juvenile myoclonic epilepsy; VPA, sodium valproate; TLE, temporal lobe epilepsy; CI, confidence interval; FWE, family-wise error | | | |

| **eTable 2:** *Clusters that survived familywise-error correction in effect of VPA dose for cortical thickness* | | | |
| --- | --- | --- | --- |
| ***JME*** | | | |
| **Cluster** | **T statistic** | ***p***_FWE_ | **Regions** |
| 1 |  | <.0001 | Left precentral gyrus  Left paracentral gyrus  Left superior postcentral gyrus  Left medial superior frontal gyrus  Left precuneus |
| 2 |  | <.0001 | Right precentral gyrus  Right supramarginal gyrus  Right paracentral gyrus  Right precuneus  Right superior parietal lobule  Right inferior parietal lobule |
| ***TLE:*** *no clusters survive familywise error correction* | | | |
| Abbreviations: JME, juvenile myoclonic epilepsy; VPA, sodium valproate; TLE, temporal lobe epilepsy; CI, confidence interval; FWE, family-wise error | | | |
